# The Educational pilot of Orchid: a new digital health intervention for reproductive life planning and education

**DOI:** 10.64898/2025.12.04.25341624

**Authors:** Maitri Shila Tursini, Catherine Stewart, Helen Carr, Alice Howe, Jennifer Hall

**Affiliations:** Reproductive Health, Institute for Women’s Health, University College London, London, UK; NHS Surrey Heartlands Health and Care Partnership, Guildford, Surrey, UK

## Abstract

**Background:** The College of Sexual and Reproductive Health’s (CoSRH) Hatfield Vision underscores the need for reliable sexual and reproductive health education and outlines specific actions to support teachers in delivering it. Orchid is a novel digital health intervention designed to empower users to understand and manage their reproductive lives, while also serving as an educational aid. This study piloted Orchid in a secondary school setting to assess its feasibility and acceptability as a tool to support the relationships and sex education (RSE) curriculum.

**Methods:** Orchid was created with members of our co-development group. We carried out three in-person sessions at a secondary school: 1) students used Orchid, working through pre-set scenarios, 2) Student focus groups, 3) a careers session. Two teachers were interviewed separately. The data were analysed using thematic analysis in NVIVO 14.

**Results:** Orchid was found to be acceptable and valuable intervention. Existing literature highlights that RSE often prioritises biological content over practical, reproducible learning, and Orchid was seen as a constructive response to these gaps. Pre-set scenarios were useful for exploring the full reproductive life cycle. Teachers and students highlighted that the digital format enhanced engagement and contributed to the development of students’ technological competencies.

**Conclusion:** Our study showed that Orchid was well received as a tool for supporting the RSE curriculum in secondary schools. Wider implementation of digital health educational tools would support delivery of the CoSRH Hatfield Vision. Further work is required to develop the content of Orchid and understand it’s potential impact.

## Introduction

Holistic relationships and sex education (RSE) aims to empower young people to make healthy, conscious, and satisfying choices regarding relationships, sexuality, and emotional and physical well-being.[1, 2] Good RSE has a positive impact on power dynamics in relationships,[1] delays initiation of sexual activity, reduces teenage pregnancy, abortions and STI rates, and increases usage of condoms and other contraceptives. [3] Receiving RSE in school has proved to be the most effective method. [4, 5]

The Hatfield Vision, developed by the College of Sexual and Reproductive Health (CoSRH), highlights the urgent need for accessible and effective RSE. It advocates supporting educators in delivering a practical, evidence-based RSE curriculum effectively.[4] In 2025, the UK Department for Education issued guidance emphasising the importance of empowering young people to take control of their health. These guidelines call for schools to integrate reliable, sensitive, and externally supported resources that address the full reproductive life course, including fertility, menopause, menstrual health, and common gynaecological conditions. [5]

Current research indicates that RSE in the UK falls short of meeting young people’s expectations and needs; [6] the curriculum tends to focus heavily on biological aspects, neglecting the practical, emotional, and social dimensions of reproductive health.[6, 7] Moreover, topics, such as fertility, preconception health and LGBTQ+ inclusion, are inconsistently covered or completely absent. [3, 6, 8, 9] Given that almost half of all UK pregnancies are unplanned, [10] that abortion rates are the highest they have ever been, [11] and that contraception use is falling,[12] providing informative, user-friendly RSE to young people is an immediate priority.

In response to these challenges, we co-developed ‘Orchid’, following the intervention mapping process.[13] Orchid is a novel digital health intervention designed to support users in developing their own reproductive life plan and learn about their reproductive health. Orchid was created with a co-development group of end users to ensure its content is user-informed and relevant.[14] Upon accessing Orchid, female users answer a set of questions known as the Desire to Avoid Pregnancy scale (DAP) to assess their pregnancy preference and calculate their likelihood of pregnancy within the next 12 months.[15] All users are then guided down a dynamic pathway of questions to formulate a customised reproductive life plan (RLP) with reliable, tailored resources. Orchid also includes evidence-based resources on common reproductive health conditions. This paper describes our pilot study in a secondary school to understand the feasibility and acceptability of Orchid as an educational support tool.

## Methods

We contacted secondary schools in London via public registry and personal networks, recruiting a state school in South London. After necessary discussions and approval with school authorities, the Orchid pilot was implemented during an enrichment programme for Year 12 students (age 16-17) interested in health sciences.

Information sheets were provided to students and their families a month before the pilot. The class teacher obtained written informed consent before the first session; students could opt out, but none did. Students explored Orchid in class using pre-set scenarios that covered different stages of the reproductive life course, including topics related to contraception, consent, menstrual health, STIs, preconception health, cancer screening, and menopause. At the end of the session, students completed an evaluation of Orchid. The following week, we conducted focus groups with students using co-developed topic guides. Based on teacher input and student preferences, students were grouped by sex and could opt for audio or video recording. Two teachers who observed the first session were later interviewed via Zoom. Teachers and students received a £25 gift voucher for their participation in an interview or focus group.

### Data Analysis

Recordings were transcribed, anonymised, and the original files deleted. Thematic analysis was conducted in NVIVO 14 by one main researcher and reviewed by a second. Themes were derived deductively from topic guides and inductively from discussions.

### Patient and Public Involvement

Orchid was co-developed from inception with 21 public contributors. The concept emerged from prior work with members of the public, highlighting the need for a trustworthy digital tool for reproductive life planning. The co-development group reviewed design, content, and study materials via regular surveys and quarterly meetings.

The pilot received ethics approval from UCL, reference 3974/005.

## Results

Fifteen students used Orchid during a lesson. Three focus groups were conducted: one video-recorded with seven female students, and two audio-recorded; one with eight female students and one with two male students.

### Content

The female focus groups stressed the need for consistent, accessible resources due to variability in education: *“I think, having it [Orchid] as a resource … is really important, because people learn differently and like the education is different everywhere*” (female student A5). One student cited only being taught about the emergency pill, whilst another reported only being taught about short-acting contraceptive methods, and several had never heard of the contraceptive patch.

Teachers noted that most RSE education takes place in either science or Personal, Social, Health and Economic (PHSE) classes, the latter are taught by staff with varied backgrounds, leading to inconsistent coverage and “*some limitations to their knowledge*” (teacher 2). Orchid was seen as a helpful external tool to provide structure and reproducibility for lessons.

### Accessibility

Orchid was generally perceived as accessible with graspable language. However, some students found terms like ‘antenatal’ and ‘premenstrual syndrome’ unfamiliar. Some felt the DAP questionnaire at the start to be lengthy: “*I would have to answer a lot of questions, that kind I felt a bit reluctant to do it*” (female student A4).

### Relevance

Overall, students felt that Orchid was relevant and provided practical insights into reproductive health that went beyond the standard biology curriculum. As one female student reflected, *“Before [this pilot], I feel like we knew the names [of contraceptives]* … *but we didn’t actually learn which one would fit for you and your lifestyle* …*you just memorised [the names]*.*”* (female student V1). Teachers shared similar views, noting that while topics such as menstruation and menopause are covered in class, the teaching often remains quite restricted: “*They are taught about menstruation, they’re taught about the menopause… they’re taught about six glands, and they’re taught basically to talk about LH / FSH and then also oestrogen and progesterone*… *And they’re… they’re told to learn it as facts rather than understanding what is actually going on*” (teacher 2).

Students appreciated that Orchid offered exposure to aspects of reproductive health they would not normally encounter, particularly through the use of pre-set scenarios. Some students found the scenarios that reflected their current stage of life the most relatable, whereas those based on future life circumstances were harder to engage with. Nonetheless, many valued these as opportunities to think ahead and understand experiences beyond their own.

Discussions also revealed differences in how male and female students perceived the program. Several male students felt that Orchid covered information they already knew, for example, “*how to check for cancer… and those kinds of prevention like contraception, I feel like that’s just taught in school. So, it wasn’t really that helpful*” (male student 1). In contrast, many female students emphasised the value of both boys and girls learning about issues that affect one another, noting that this promotes greater mutual understanding. They acknowledged that they often lacked awareness of the male perspective and believed this was a reciprocal gap. For example, some pointed out that topics such as emergency contraception were not well understood by male peers, who tended to underestimate its side effects and implications.

Overall, Orchid was seen as a valuable and relevant program that encouraged students to think more broadly about reproductive health, not just as biological facts, but as real-life issues that affect everyone differently.

### Feasibility & Integration

Teachers and students agreed that integrating Orchid into the RSE curriculum would be feasible and beneficial for learning. However, teachers reported challenges accessing RSE content on school networks due to restrictive firewalls. Despite this, they saw potential for Orchid to be used in class with appropriate collaboration with the school’s IT department or as homework. Digital tools were considered engaging “*because I think it’s so different from all structured lessons, we’d probably pay attention to it a bit more*” (female student, A5) and teachers noted the broader educational value of digital integration: “*Considering the times we live in now, digital and IT use in education is very important*” (teacher 1).

Regarding session logistics, most students preferred a combination of mixed-gender and gender-separated sessions. While they acknowledged that some topics might be uncomfortable in mixed groups, they emphasised the importance of all students learning about key topics such as menstruation, noting that boys often lacked this knowledge.

Students also suggested that topics like menstrual health should be introduced earlier, ideally in primary school, to ensure understanding before menarche. Given that menarche is occurring at younger ages, early coverage was viewed as essential. Other topics could be introduced progressively, for example, reproduction in Year 6 (ages 11–12) and contraception in Year 8 (ages 13–14).

Teachers indicated that, with appropriate training and the inclusion of an effective feedback mechanism to address issues, sustained use of Orchid would be feasible.

## Discussion

This pilot study showed that it is feasible and acceptable to use Orchid as an educational tool in a secondary school setting. Students and teachers found it engaging and relevant, and they thought it aligned well with current educational needs, echoing the priorities of the Hatfield Vision, which advocates for accessible, evidence-based digital health resources.[4]

One of the key themes that emerged was inconsistent RSE teaching that focuses on biology rather than on practical learning, a theme widely recognised in the literature.[3, 16] Orchid was seen as a positive solution to this. The use of pre-set scenarios was well received, offering a structured way to explore RSE topics and aligning with previous calls to convey information engagingly, enabling young people to integrate it into their current life stage. [17] The digital format was seen as a strength, reflecting the broader trends in digital education.[18]

## Data Availability

All data produced in the present study are available upon reasonable request to the authors

## Implications for Practice and Policy

This study supports integrating co-developed digital health interventions, such as Orchid, into RSE curricula to reduce reliance on informal sources and ensure consistent, accurate information. As healthcare continues to embrace digital innovation, tools like Orchid offer a potential way to expand access to essential RSE knowledge.

Policymakers and educators should invest in scalable, evidence-based resources that complement existing teaching and support staff in delivering RSE content.

## Future Research

Further research should evaluate Orchid’s long-term impact on reproductive health knowledge, attitudes, and behaviours. Exploring effectiveness across diverse school settings will be key to ensuring equity and accessibility.

